# Ambulatory vEEG discontinued against medical advice among individuals with intellectual disabilities: a retrospective chart review

**DOI:** 10.1101/2023.11.26.23299038

**Authors:** Ewan S. Nurse, Nicholas Winterling, Mark J. Cook

**Affiliations:** Seer Medical, Melbourne, 3000, Australia; Department of Medicine, St. Vincent’s Hospital Melbourne, The University of Melbourne, Fitzroy 3065, Australia; Department of Biomedical Engineering, The University of Melbourne, Parkville 3052, Australia

**Keywords:** ambulatory video EEG, epilepsy, intellectual disability, discharge against medical advice

## Abstract

**Objective:** This retrospective chart review aims to quantify the rate of patients with intellectual disability (ID) accessing an Australian ambulatory EEG service, and understand the implications of discontinuing studies against medical advice.

**Methods:** Electronic records of referrals, patient monitoring notes, and EEG reports were accessed retrospectively. Each referral was assessed to determine whether the patient had an ID. For each study where patients were discharged against medical advice, the outcomes of their EEG report were assessed and compared between the ID and non-ID groups. Exploratory analysis was performed assessing the effects of age, the percentage of the requested monitoring undertaken, and outcome rates as a function of monitoring duration.

**Results:** There were significantly more patients in the ID group with early disconnection than the non-ID group (Chi squared test, *p*=0.000). There was no significant difference in the rates of clinical outcomes between the ID and non-ID groups amongst patients who disconnected early.

**Conclusions:** Although rates of early disconnection are higher in those with ID, study outcomes are largely similar between patients with and without ID in this retrospective analysis of an ambulatory EEG service.

**Significance:** Ambulatory EEG is a viable alternative to inpatient monitoring for patients with ID.

**Highlights three bullet points (one sentence each, with a maximum of 130 characters - including spaces:**

- Recording EEG remains a challenge for patients with epilepsy and ID
- This study shows that patients with ID are more likely to disconnect early compared to those without ID
- Study outcomes are not significantly different between those with and without ID, however some outcomes may be underpowered

## 1. Introduction

Epilepsy occurs in approximately one in four people with intellectual disabilities (IDs) (Kerr et al., 2014). Although prolonged EEG is considered useful in the diagnosis of an epilepsy in people with ID, it is recognised that undertaking an EEG presents unique challenges in this population across both patient-specific and systemic factors (Devinsky et al., 2015; Gotlieb et al., 2023).

Improving access to EEG, and the tolerability of the testing, has been considered a priority in the management of epilepsy in patients with ID (Kerr et al., 2009). Even amongst the broader epilepsy population, there is increasing awareness of the importance of patient comfort during EEG assessment (Egger-Rainer et al., 2022). Amongst the broader population of patients receiving inpatient neurological care, patients with epilepsy have the highest rates of discharge against medical advice of approximately 2-3%, with risk factors including young age, low socioeconomic status, and drug/alcohol use, however these factors are by no means specific to the epilepsies (Agarwal et al., 2021; Raja et al., 2020).

Interestingly, these works have not identified intellectual disability as a risk factor for premature discharge against medical advice from epilepsy monitoring units, despite concerns amongst the neurology community around the tolerability of EEG (Devinsky et al., 2015). This omission may be due to potential lack of representation of epilepsy and ID in the literature (Kerr et al., 2014; Kinney et al., 2020; Shankar et al., 2018).

Ambulatory EEG may provide a favourable alternative to inpatient monitoring for people with ID, as many of the potential stressors of the hospital environment are avoided (Kandler et al., 2017). A recent review of at-home EEG technologies for adults with ID by Mline-Ives et. al concluded that ambulatory EEG was a feasible solution for monitoring people with ID, but that more research is needed to quantify the utility (Milne-Ives et al., 2023).

This work presents a retrospective chart review of an Australian ambulatory video EEG service, assessing the rates of early study secession in patients with and without ID. Clinical outcome measures such as rate of abnormal EEG capture and rate of clinical event capture are compared between the ID and non-ID groups of the patients who completed their studies early. Sub-analyses assessing the effects of age and duration of monitoring were undertaken.

## 2. Methods

### 2.1 Participants

This study was conducted under approval from the Human Research Ethics Committee of St. Vincent’s Hospital Melbourne (042/18). All participants provided written, informed consent. Clinical and patient support records of all patients undertaking ambulatory video EEG (vEEG) from October 2022 to September 2023 at an Australian service (Seer Medical Pty. Ltd) were accessed retrospectively. All records were stored in a secure cloud platform.

Each participant was referred for ambulatory video EEG, conforming to Australian Medicare guidelines. Typical referral questions were for differential diagnosis, querying ongoing seizure activity, or classifying seizure types.

Patients attended a clinic for the fitting of the device. EEG is recorded with the standard 10-20 system at 250 Hz and 24 bits per sample. Three ECG contacts are used to record heart activity. The EEG and ECG are time-synchronised to a 30 frame per second camera that has an automatically activating night-vision mode in low-light settings (Nurse et al., 2023). EEG electrodes were attached to the scalp with a novel, water-soluble adhesive allowing for 7 day recordings, and patients (or carers) to fix electrodes without a technician on site (Nurse et al., 2022). As recordings are typically undertaken without direct medical supervision, no activation methods are used to provoke seizure activity due to safety concerns. At the end of the monitoring period, patients are able to remove the electrodes by applying warm water (such as by having a shower).

### 2.2 Data collection

As part of standard clinical operations, it was noted whenever a patient prematurely stops the study against medical advice, which we define as an ‘early disconnection’ in this work. Common reasons for early disconnection were: general discomfort, difficulty sleeping, or otherwise not wishing to continue with the study. Referrals were assessed by the authors for whether the patient had an intellectual disability (ID). This was often explicitly stated, but sometimes inferred (i.e. if the patient had a prior diagnosis of Lennox-Gastaut Syndrome). The age and gender of the patient were extracted from the referral information.

The concluding report in studies where early disconnection occurred were accessed, and analysed for clinical outcomes, including: whether an event was reported by the patient, whether an event was discovered on review of the vEEG, whether the EEG was found to be abnormal, and if so, whether the abnormality was focal or generalised. If there was a focal finding, it was noted if the foci were temporal or extratemporal. An event was defined as an episode of clinical relevance to the referral question, with or without electrographic correlate. All clinical reports were written by consultant neurologists, in consultation with senior neurophysiologists.

### 2.3 Data analysis

The percentage of studies with and without ID in the overall and early disconnect groups were calculated. The gender ratio and age distributions were also compared between the ID and non-ID groups. When statistical significance tests were undertaken for the primary outcome measures, the corresponding Bayes factor analysis was also undertaken to measure if the null hypothesis could be accepted (Kass and Raftery, 1995). Of the studies in the early disconnection group, the percentage of studies with each of the clinical outcomes described in section 2.2 Data collection was assessed.

The recorded duration was assessed as a function of patient age for the ID and non-ID groups, and modelled with linear ordinary least squares regression.

Early disconnection studies were analysed for what percentage of the ordered study duration were captured, and this was compared between the ID and non-ID groups.

For each of the main report outcome measures, the duration of monitoring was compared between where there was or was not a clinical finding for that measure.

When calculating the percentages of outcome measures, a 95% confidence interval was calculated using an asymptotic normal approximation (Python statsmodels version 0.15.0). Due to non-normality of the variables being assessed, distributions were compared using the Mann-Whitney U test (Python scipy version 1.11.3). Linear regression was undertaken using ordinary least squares (Python statsmodels version 0.15.0). Bayes factor analysis was undertaken using the BayesFactor library for the Chi-square tests, and a previously published package for non-parametric Bayes factor calculation for the Mann-Whitney U tests (van Doorn et al., 2020). 5e3 samples were used to calculate the posterior samples.

Analyses were performed in Python 3.11.5 or R 4.3.1.

## 3. Results

### 3.1 vEEG study outcomes

A total of 2992 studies were captured, of which 181 (6.0%) had an identified intellectual disability (ID) in the referral. The referred duration of monitoring for the ID and non-ID groups is shown in Supplementary Table 1. The distributions are significantly different as measured by the Mann-Whitney U test (*p*=0.006), with the duration typically shorter for the ID group.

219 of the captured studies had an early disconnection noted on their record (7.3% of all studies), with 36 of these studies occurring in patients with an ID (16.4%). There were significantly more patients in the ID group with early disconnection than the non-ID group (Chi squared test, *p*=0.000), and this result is confirmed with the high Bayes factor value (1284). In the early disconnection group, 151 patients (69%) were female. There was no significant difference between the ID and non-ID groups for proportions of females (Chi squared test, *p*=0.56), however the Bayes factor of above 10 indicates there is strong evidence that females are more strongly represented in the non-ID group. The median age of the early disconnection cohort was 27 years, and there was a significant difference between the ID and non-ID groups (Mann-Whitney U test, *p*=0.007), however the Bayes factor is low, indicating this data is potentially undersampling the true distribution. The age breakdown of early disconnection patients is shown in Supplementary Figure 2.

Details of the vEEG study outcomes are shown in Table 2. None of the outcome measures were found to be significantly different between the ID and non-ID groups (Chi squared test). Although the percentage for most measures is higher in the ID group, the confidence intervals in both groups are relatively large. Hence, regardless of whether a patient has an ID, outcome measures of vEEG are statistically similar amongst those who disconnect early. The Baysian analysis however provides discordant information, demonstrating strong evidence for patients with ID to have different probabilities of having discovered events, an abnormal EEG to be focal, for a focal epilepsy to be temporal, or for an abnormal EEG to be generalised. This discrepancy is likely due to the analysis being underpowered given the effect size (Kass and Raftery, 1995)

**Table 1:** Study details.

|  | **Total cohort** | **ID group** | **Non-ID group** |
| --- | --- | --- | --- |
| **Total studies identified - count** | 2992 | 181<br>(6.0%) | 2811<br>(94.0%) |
| **Total studies with early disconnection - count (percent)** | 219 (7.3% of total identified studies) | 36<br>(16.4%) | 183<br>(83.6%) |
| **Chi squared comparison of percent ID patients - *p* value** | **0.000\*** | - | - |
| **Bayes Factor** | **1284** |  |  |
| **Female patients with early disconnection count (percent)** | 151 (68.9%) | 22<br>(64.7%) | 129<br>(69.7%) |
| **Chi squared comparison of percent female patients - *p* value** | 0.56 | - | - |
| **Bayes Factor** | **13.9** |  |  |
| **Age of patients with early disconnection - median (min, max) years** | 27 (4,94) | 20 (6, 55) | 28 (4, 94) |
| **Mann-Whitney U comparison of age distributions- *p* value** | **0.007\*** | - | - |
| **Bayes Factor** | 0.54 |  |  |

**Table 2:** Early disconnection study outcomes for ID and non-ID groups.

| **Outcome** | **ID: Count, Percent (95% CI)** | **Non-ID: Count, Percent (95% CI)** | **p-value (Chi Squared test)** | **Bayes Factor** |
| --- | --- | --- | --- | --- |
| Prior diagnosis of epilepsy | 19, 52.8% (36.5-69.1%) | 79, 43.2% (36.0-50.3%) | 0.29 | -0.619 |
| Reported events | 21, 58.3% (42.2-74.4%) | 94, 51.4% (44.1-58.6%) | 0.44 | -1.52 |
| Discovered events | 4, 11.8% (0.9-22.6%) | 18, 9.7% (5.5-14.0%) | 0.72 | **76.2** |
| Abnormal EEG | 19, 52.8% (36.5-69.1%) | 74, 40.4% (33.3-47.5%) | 0.17 | 0.628 |
| Abnormal EEG with finding of focal epilepsy | 14, 73.7% (53.9-93.5%) | 56, 75.7% (65.9-85.5%) | 0.86 | **10.2** |
| Focal epilepsies with temporal foci | 12, 85.7% (67.4-100%) | 48, 85.7% (76.5-94.9%) | 1.0 | **16.8** |
| Abnormal EEG with finding of generalised epilepsy | 5, 26.3% (6.5-46.1%) | 19, 25.7% (15.7-35.6%) | 0.96 | **9.22** |

### 3.2 Exploratory analysis

#### 3.2.1 Effect of age

To assess the effect of age on duration of monitoring captured, ordinary least squares linear regression was performed in both the ID and non-ID groups. This is shown in Figure 1. The fit to the non-ID group was not significant (*p*=0.27). The linear regression for the ID group was significant (*p*=0.03), however the coefficient of determination was poor (R^2^ = 0.13). The gradient of the fit was 1.2 hours of recording per year of age, demonstrating that older patients with ID tend to tolerate vEEG for a longer amount of time. It should be noted that the highest age in the ID group was only 55 years of age, so the study population, and hence the model, may not be representative of the broader population

**Figure 1:**
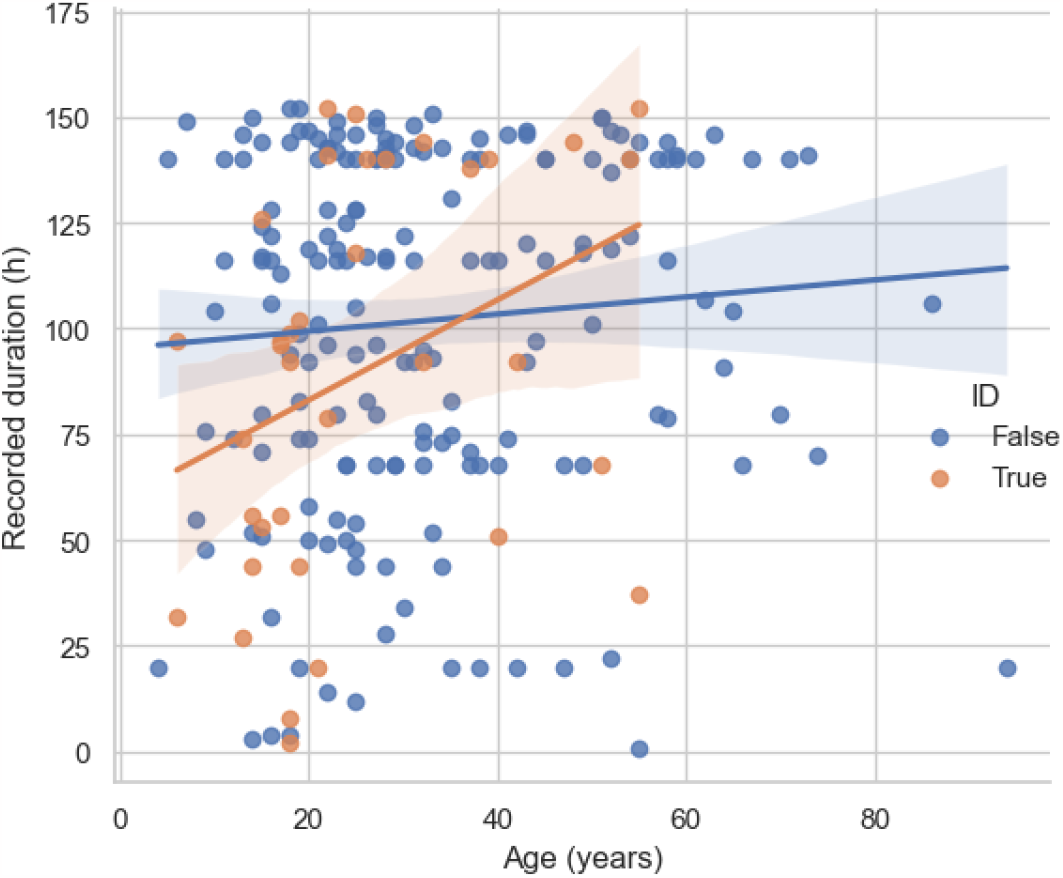
Scatterplot of study recorded duration as a function of age, with a linear fit to each of the ID and non-ID groups. The fit to the ID group (orange) is statistically significant (*p* = 0.03), however the coefficient of determination is poor (R^2^ = 0.13). The gradient is 1.2 hours of recording per year of age. The fit to the non-ID group was not significant (*p* = 0.27).

#### 3.2.2 Percentage of study captured

For each study, the percentage of the ordered duration that was able to be recorded is shown in Figure 2a, and the survival curve (i.e. a complementary cumulative density function) in Figure 2b (noting most studies are ordered for 7 days as shown in Supplementary Table 1). There is no significant difference in the distributions between the ID and non-ID cohorts (Mann-Whitney U test, p=0.49). It can be seen that approximately 40% of studies ceased after >80% of the ordered study duration had elapsed. Although there is some divergence between the distributions after approximately 2 days, they appear to reconverge at approximately 3 days.

**Figure 2:**
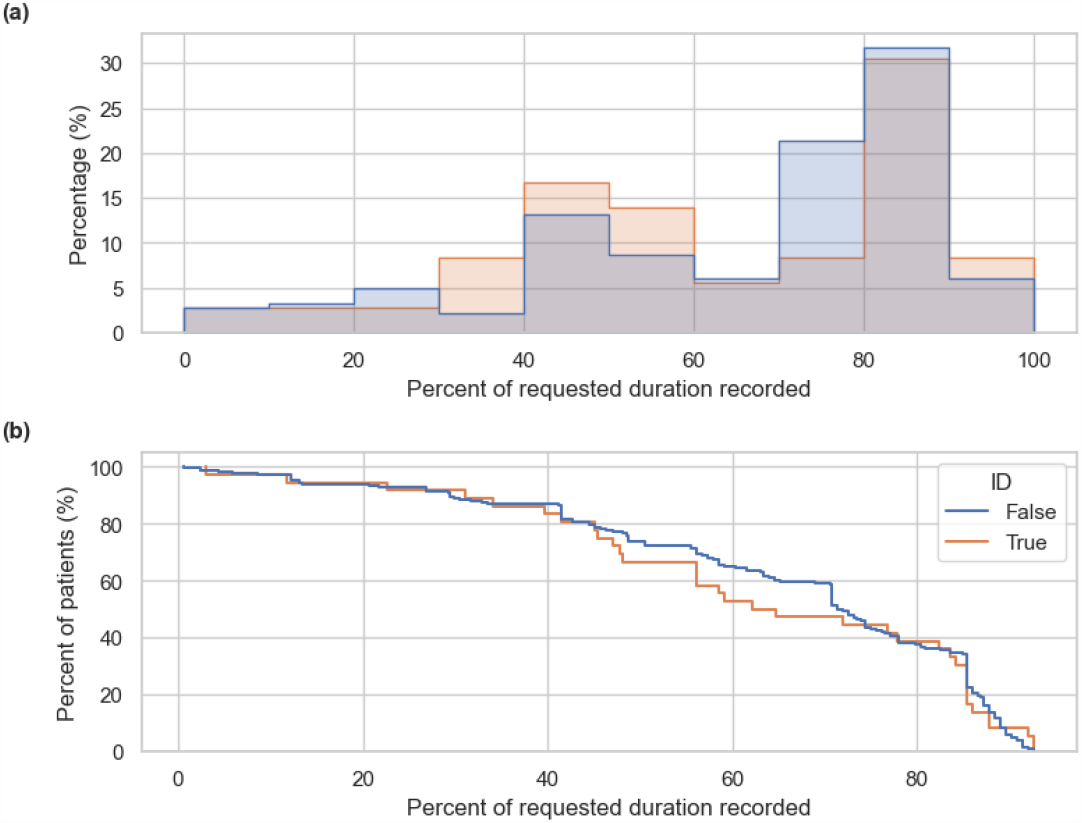
**(a)** Histogram of recorded durations represented as the percentage of requested study duration, and **(b)** survival curve (complementary cumulative density function) of recording duration. The distribution of the percentage of requested duration recorded was not significantly different between the ID and non-ID groups (Mann-Whitney U test, p=0.49).

#### 3.2.2 Clinical outcomes as a function of recorded duration

For each of the primary clinical measures listed in Table 2, we examined the difference in recorded duration. Figure 3 shows boxplots of the distributions for each outcome. None of the groups were significantly different (Mann-Whitney U test). Prior epilepsy diagnosis and abnormal EEG had a higher median recording duration, perhaps due to previous experience with EEG recordings. Patients who had reported and discovered events had shorter median durations, perhaps suggestive of patient frustration with ongoing recording after having an event, which is consistent with the previous literature (Agarwal et al., 2021; Raja et al., 2020).

**Figure 3:**
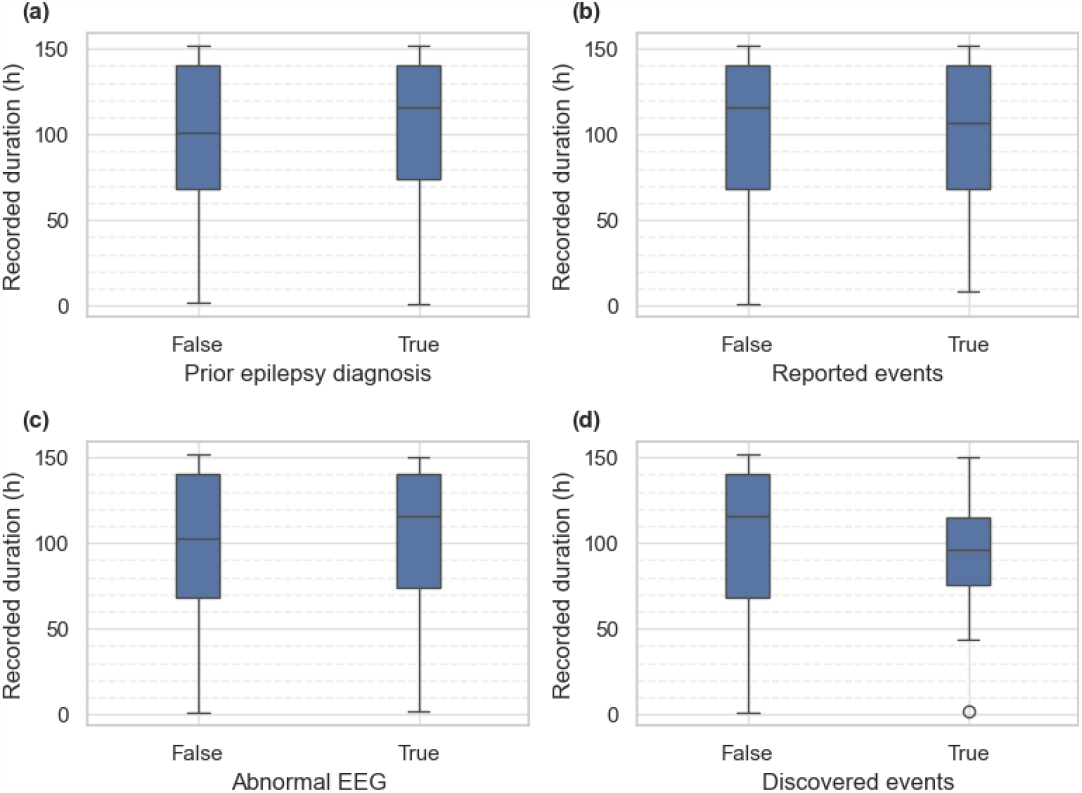
Boxplots of recording duration comparing clinical outcomes. None of the pairs are significantly different (Mann-Whitney U test).

## 4. Discussion

This study reports on the rate of ID in patients referred to an Australian ambulatory video EEG service, and the effects of early disconnection on study outcomes.

Results from this work demonstrate that rates of early study cessation seem higher than those of inpatient services (7% compared to 2-3%). This is perhaps to be expected, as the lack of direct clinical supervision in ambulatory monitoring may put less pressure on patients to persist with long-term EEG recordings. There is also potential that patients accessing at-home monitoring would not have presented for inpatient monitoring, and although their recording was not as long as the requested duration, multiple days of EEG were captured in the majority of patients (median of 113h). Furthermore, the adhesives used in this service are water soluble, hence patients are able to remove the equipment without the use of solvents.

The significant, positive, linear trend of age with recording duration in the ID group may indicate that shorter durations of monitoring should be ordered for younger patients with a known ID. Furthermore, many patients having epileptic seizures tend to have an event within the first 3 days of monitoring (Wong et al., 2023). However, due to the heterogeneity of this cohort, further research is required to understand if specific causes or severity of ID are associated with less tolerance for EEG monitoring.

Discontinuing EEG monitoring in an ambulatory setting may present a safer set of circumstances than inpatient monitoring units, as patients will not have undergone any deliberate seizure provocation methods, and hence are not in a heightened state of seizure risk, or do not need to re-titrate to their standard medication levels before returning to a baseline level of seizure risk (Rheims and Ryvlin, 2014). Hence, rates of early disconnection may be higher due to the decreased risk of seizure clusters or status epilepticus.

The results presented in this work, to our knowledge, are the first to describe discharge against medical advice from ambulatory EEG monitoring, in both ID and non-ID cohorts. The total analysed cohort is relatively large, assessing nearly 3000 records collected from over 20 clinical sites across Australia. However, even with this relatively large starting cohort, some of the analyses were still likely underpowered as measured by Baysian analysis. This Baysian analysis was useful in elucidating this point, that would not have been demonstrated by frequentist analysis alone (Leong et al., 2019). We encourage other services to undertake similar reviews in order to better understand service provision to patients with ID.

This work has several limitations. Identifying ID required the information to be provided in the referral, which may not always be the case. Furthermore, the existence of an ID may not be known to the referring clinician. Data were only collected from a single ambulatory service provider, and may not be representative of the broader epilepsy and ID populations. Ideally, prospective studies would explicitly collect information about ID from referrers, carers, and patients. Although the initial number of assessed studies is large, the number of patients in the ID group who disconnected early is relatively small (n=36), and hence important sub-analyses such as understanding outcomes for specific epilepsy syndromes or other ID causes cannot be undertaken. Similarly, there are not enough records to understand demographic or socioeconomic factors that might contribute to early disconnection in the ambulatory setting.

Having epilepsy and ID has a profound impact on the lives of people with these conditions and those who care for them. Hence, the provision of EEG to this population is of significant importance (Kerr et al., 2014). This work has quantified the rates of patients with ID accessing an Australian ambulatory EEG service, and assessed the rates of, and implications of, discontinuing studies against medical advice. A small, but significant positive correlation between age and duration of monitoring was found in those with ID. We have demonstrated that although patients with ID are over-represented amongst patients stopping their study early, the clinical outcomes remained the same as those without ID. Furthermore, the distribution of the percentage of the requested duration that was recorded is the same between the ID and non-ID groups. Recording in the home for long durations ensures that even with premature study cessation, sufficient information is regularly still captured to answer the referring clinical questions.

## Data Availability

All data produced in the present study are available upon reasonable request to the authors

## Contributions

Conceptualization: E.S.N.; Formal analysis: E.S.N and N.W.; Methodology: E.S.N, N.W. and M.J.C.; Software: E.S.N and N.W.; Supervision: M.J.C.; Writing – original draft: E.S.N. and M.J.C.; Writing - review & editing: E.S.N. and N.W. and M.J.C. All authors approved the final article.

## Supplementary

**Supplementary Table 1:** Count of referral durations for ID and non-ID groups. The distributions are significantly different as measured by the Mann-Whitney U test (*p*=0.006)

| Duration (days) | ID group (Count) | Non-ID group (count) |
| --- | --- | --- |
| 3 | 7 | 11 |
| 4 | 1 | 10 |
| 5 | 2 | 1 |
| 6 | 0 | 2 |
| 7 | 25 | 159 |

**Supplementary Figure 2:**
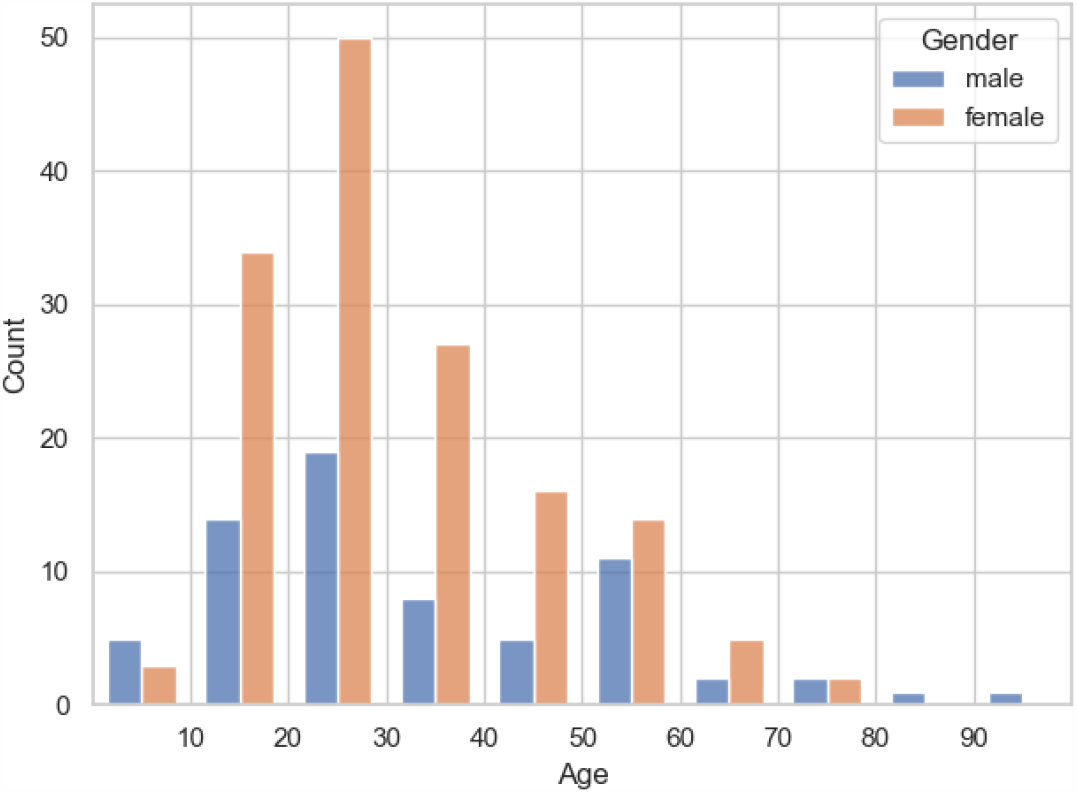
Age breakdown of early disconnection patients, separated by gender.

